# Exploring stroke survivors’ and physiotherapists’ perspectives of the potential for markerless motion capture technology in community rehabilitation

**DOI:** 10.1101/2022.03.18.22272596

**Authors:** Alice Faux-Nightingale, Fraser Philp, Enza Leone, Brinton Boreman Helliwell, Anand Pandyan

## Abstract

**INTRODUCTION:** Many stroke survivors do not receive optimal levels of personalised therapy to support their recovery. Use of technology stroke rehabilitation has increased in recent years to help minimise gaps in service provision. Markerless motion capture technology is currently being used for musculoskeletal and occupational health screening and could offer a means to provide personalised guidance to stroke survivors struggling to access rehabilitation.

**AIMS:** This study considered context, stakeholders, and key uncertainties surrounding the use of markerless motion capture technology in community stroke rehabilitation from the perspectives of stroke survivors and physiotherapists with a view to adapting an existing intervention in a new context.

**METHODS:** Three focus groups were conducted with eight stroke survivors and five therapists. Data were analysed using reflexive thematic analysis.

**RESULTS:** Five themes were identified: limited access to community care; personal motivation; pandemic changed rehabilitation practice; perceptions of technology; and role of markerless technology for providing feedback.

**CONCLUSIONS:** Participants identified problems associated with the access of community stroke rehabilitation, exacerbated by Covid-19 restrictions. Participants were positive about the potential for the use of markerless motion capture technology as a means to support personalised, effective stroke rehabilitation in the future, providing it is developed to meet stroke survivor specific needs.

## Introduction

Stroke is a leading cause of long-term disability, often resulting in a combination of sensory-motor, communication, visual, and cognitive impairments[1]. While some survivors recover, many remain with considerable levels of disability after stroke[1], and these disabilities can affect stroke survivors’ quality of life, and limit their ability to reintegrate into society or return to employment.

In the UK, existing guidelines recommend admission to a hyperacute stroke ward as soon as possible, where, depending on the mechanism, people affected by stroke will receive the necessary imaging, monitoring and thrombolytic or antithrombotic management[2]. Early mobilisation, forming the initial basis of stroke survivors physical therapy rehabilitation journey, is recommended within 24 hours of the onset of the stroke[2]. Whilst guidelines recommend that access to rehabilitation services should be determined by stroke specific goals, access and onward referral to further inpatient rehabilitation, specialist stroke therapy rehabilitation centres and early supported discharge services vary according to availability, length of stay, or number of sessions provided. Rehabilitation, which aims to minimise activity limitations and participation restrictions, may draw on repetitive, task-specific practice with appropriate equipment and feedback in a functionally relevant context. Appropriate levels of rehabilitation, which integrate these principles, can lead to improved movement and outcomes[3-5], while restricted rehabilitation access is associated with poor recovery profiles and readmissions[4]. Recently updated guidelines recognise the link between increased access to rehabilitation and improved recovery[2]. The recommended dosage in the new guidelines has increased from 45 minutes a day, seven days a week, to 3 hours of multidisciplinary therapy a day at least 5 days a week[2]. However, it is acknowledged that previously recommended volume of therapy was commonly inaccessible for patients[6-8] and stroke survivors can experience difficulties in adequately accessing rehabilitation services following discharge[6, 9-13]. Research suggests that home-based rehabilitation is effective[14] and promotes positive clinical outcomes, and that this can lead to greater satisfaction amongst stroke survivors, reduced caregiver strain, and reduced hospital readmission rates and length of stay[15].

Telerehabilitation, the delivery of rehabilitation using technology, and use of remote monitoring sensors can be beneficial for patients due to their ability to support access to services where clients face limitations to attending in person, e.g. in cases of geographical isolation[16]. Telerehabilitation facilitates patients as they engage with rehabilitation in their own time and space[17], and has been suggested to promote engagement with rehabilitation practices[18]. These services can offer the most benefit where they include access to information and feedback about lifestyle, risk factor modification, and therapy for addressing the impairments resultant from stroke. In stroke rehabilitation, telerehabilitation is currently used to support areas like mobility, speech, and cognition, and can build stroke survivors’ confidence with these activities [18]. Current advances, such as the development of markerless motion capture technology, which uses a camera to measure the ability to move or carry out functional tasks, could further support remotely delivered telerehabiltation for motor recovery and the physical effects of stroke. Single camera, markerless systems are easy to setup, require no specialist skill for the capture and interpretation of data compared to hospital or laboratory systems so can be used in patients’ homes, overcoming access barriers associated with hospital-based services[19]. However, it is important that implementation of technology into services does not further embed social health inequalities and is acceptable and feasible for the intended users and providers of the service [20].

The NHS long-term plan sets out the requirement for a new service model for the 21^st^ Century in which digitally enabled care is considered mainstream across the NHS[21]. These principles are reflected in the recent guidelines which recognise that technology could be used to augment existing delivery of rehabilitation stroke services, expedited by the pandemic. Integration of technology into rehabilitation practice can extend the ability of patients to access this support and guidance remotely compared to conventional rehabilitation models in which guidance for an individual’s rehabilitation comes from in-person sessions with rehabilitation professionals[18]. The Covid-19 pandemic has affected delivery of health services across the nation, including in acute and subacute stroke rehabilitation[22] and community based rehabilitation services[23]. These changes affected stroke care[24], restricting stroke survivors from accessing necessary services. As UK government policy continues to embrace digital facilities in the care pathway[25] and services are developed for digital use and delivery of complex interventions, it is important to gather evidence about the views and experiences of the intended users. Doing so ensures that the facilities meet user needs through: engagement of stakeholders, identification of key uncertainties, intervention refinement, and consideration of the overall context in which the service and intervention are positioned[14].

This study is positioned within a wider project which hopes to develop an intervention for stroke rehabilitation within the community by adapting an existing intervention (markerless motion capture) in a new context (stroke rehabilitation in the community). As part of this process, this study considered context, stakeholders, and key uncertainties in this area, exploring stroke survivor and physiotherapist attitudes towards the use of markerless motion capture technology within community stroke rehabilitation.

## Methods

This study used focus groups to explore stroke survivors’ and physiotherapists’ lived experience of stroke rehabilitation and perceptions of the use of markerless motion capture technology for stroke rehabilitation. Interpretive description methodology[26] was engaged within the study, allowing researchers to explore and record the subjective experiences of groups and use the findings to develop evidence-based knowledge to inform practice. Interpretative description is used to investigate patient experiences[27-29] and perspectives[30, 31] of illness, healthcare and healthcare environments. It uses clinical knowledge and experiences and perceptions of participants to develop descriptive findings which can be applied to clinical practice and used to inform future design[26].

Ethical approval for the study was given by the University Research Ethics Committee Review (MH-210173) prior to recruitment taking place. This paper follows the COREQ guidelines[32], checklist attached as supplementary material.

### Recruitment

UK stroke survivors and physiotherapists with experience in stroke rehabilitation were invited to attend focus groups using convenience sampling strategies. Participants were invited if they were either 1) people who had a stroke, who had experienced rehabilitation in the community, and who could communicate about their experiences or 2) physiotherapists with experience in stroke rehabilitation. Stroke survivors were provided information about the study and invited to participate using adverts on social media and a stroke specialist therapy centre (ARNI) mailing list. Therapists were invited using social media adverts and a professional society (ACPIN) mailing list.

Prospective participants were given an information sheet and consent form and could ask the researcher questions prior to giving consent. Audio recordings or speech-to-text were offered on all stroke survivor documents to increase accessibility. All participants who expressed interest in the study consented to participate in a focus group. One participant with some language difficulties asked to be accompanied by their carer who could speak on their behalf as necessary. The carer also consented to participate in the study.

### Data collection

The focus groups took place during the Covid-19 pandemic (May-June 2021) and all participants were affected by the regulations and pressures of the period. After agreeing to the arrangements for the session, four participants struggled to attend the focus group. In recognition of the circumstances, anyone who had consented to participate but who could not attend the session was offered the opportunity to discuss the focus group topic guide in an interview-style sitting (referred to as ‘interviews’ from here on) at a later point. Where this happened, the interviews followed the same topic guide as the focus groups.

Three focus groups took place, two with stroke survivors, and one with therapists, each session included up to five participants. Two interviews were carried out with therapists who couldn’t attend the focus group. Focus groups were hosted virtually, and were facilitated by two members of the research team, one clinical researcher (AP, PhD), male, with experience in qualitative research and technology to support rehabilitation, and one non-clinical researcher (AFN, MPhil), female, with experience in qualitative healthcare research. Interviews were conducted by only the non-clinical researcher. Sessions lasted 90 minutes. Conversation was directed by a topic guide and a short video of existing markerless technology currently used for musculoskeletal and occupational health screening (https://www.youtube.com/watch?v=PT2zA39xnuY). Participants were asked to talk about their experiences of post-stroke rehabilitation and their thoughts about the potential for markerless motion capture technology to support stroke rehabilitation (Appendix 1), although the topic guide was used flexibly within the session and facilitators responded to topics raised by the participants. Stroke survivors were offered breaks and were encouraged to pause if they needed to rest e.g. if they felt fatigued.

All focus groups and interviews were video and audio recorded and transcribed verbatim to produce anonymised transcripts used for analysis. AFN took notes during the focus groups.

Sample size was informed by the concept of information power, considering the study’s defined aim, focused data gathering, preparation and ongoing review [33]. Recruitment, however, was limited by events of the Covid-19 pandemic which took place during this study period.

### Data analysis

Reflexive thematic analysis was carried out following the guidelines by Braun & Clarke[34]. Codes were generated inductively for all transcripts by one non-clinical author (AFN) using open coding techniques. Codes were checked by two members of the research team, one clinical (EL) and one non-clinical (AFN), facilitated by NVivo. Themes and subthemes were generated iteratively by both researchers independently according to commonality across the dataset, significance to the participants, and relevance to the research questions. The preliminary themes and findings from each researcher were brought to a wider project meeting where they were developed and refined with regular revisiting of the data to confirm understanding and ensure that the analysis accurately represented participants’ experiences.

### Reflexivity

The research team was constructed of people from a range of experiences, including clinical academic physiotherapists, a health researcher, a stroke survivor and an academic bioengineer. A reflexive approach was taken, where the research team brought knowledge and insight from their experiences and disciplines to develop a meaningful analysis of the accounts, acknowledging the influence of their background on the focus and interpretation of the data.

## Analysis

Eight stroke survivors participated in the stroke survivor focus groups (five women, three men) with one further participant (carer) who sometimes spoke on behalf of a participant with speech impairments. Stroke survivors’ strokes ranged from 6 months to 5+ years prior to the focus groups. Five physiotherapists took part study, all women, all physiotherapists working in NHS stroke rehabilitation services with 3-20 years of experience, three in one focus group, two interviews. Roles included acute neurorehabilitation roles, early supported stroke discharge team, and community stroke services.

## Findings

Five themes were identified across the stroke survivor and therapist focus groups:

⍰ Limited access to community care

⍰ Personal motivation

⍰ Pandemic changed rehabilitation practice Ill Perceptions of technology

⍰ Role of markerless technology for providing feedback

### Limited access to community care

Stroke survivors described feeling that they were not given enough support post-discharge from hospital and described struggling to access follow up support in the community, and this was also raised by the physiotherapists.

*“I think I find it interesting that everybody in this group has said that they want to progress more. They want to do more but there aren’t the facilities*.*” 04, stroke survivor*

*“I think I think we see that patients always want to do more, we just sadly haven’t got the time*.*” Physiotherapist DH*

Limited access was presented through: long waiting lists; inability to access appropriate community services due to lack of referral, services unavailable in the area, or referral to services not specific to stroke; strict entry criteria for services; or services cut off after a fixed number of weeks. Individual stroke survivors also mentioned being unable to access support due to personal limitations:

*“I had some other problems because I can’t drive [… and I] can’t get someone to take me*.*” 01, stroke survivor*

### Personal motivation

Although stroke survivors described difficulties accessing community rehabilitation services, most expressed high levels of motivation to support themselves with their post-stroke rehabilitation.

*“I’m doing my own physio, obviously, continue on from the NHS basic physio and I’m working through [a] stroke manual” 02, stroke survivor*

Stroke survivors’ described searching for additional stroke specific programmes, community programmes, stroke specialist therapists/trainers, and research projects which could offer guidance and support. Financially able stroke survivors described attending stroke services/facilities or buying rehabilitation equipment/technology, but this was not available to everyone and many stroke survivors described limitations in the resources they could access due to cost. This encouraged some to participate in research studies which were free and facilitated access to rehabilitation training or equipment:

*“I just keep looking on the internet to see whether there’s anything that’s come along. Either technology-wise or any otherwise to see what improvements can be made or what research is going on and if there’s something that I think that might be useful to me, then I’ll join in with it*.*”* 03, stroke survivor

This participant described using existing stroke rehabilitation technology (e.g. GripAble) and said that gamified exercises and feedback on progress further improved their motivation to engage with rehabilitation:

*“[It is] a bit of fun and you measure your own progress, which is important”. 03, stroke survivor*

### Pandemic changed rehabilitation practice

All participants discussed the impact of the Covid-19 pandemic on rehabilitation services. Therapists described restrictions in community services and the associated increases in time that stroke survivors spent without access to therapist-led programmes. Stroke survivors described impact of the loss of home visits on their practice:

*“I cannot do exercises on my own but if I have somebody there to help me, it’s spurs me on to do more […] but then when the pandemic started, they said they couldn’t do it again*…” 01, stroke survivor

Stroke survivors described how social distancing regulations had caused more services to offer virtual services which inadvertently increased access to a broader geographical area. This was very well received by stroke survivors, particularly those who lived in areas with few available services.

*“[…] Since Covid-19, obviously, [in-person rehabilitation] hasn’t been able to happen, but I found lots of things online, so I actually think it’s been a positive thing that people, organizations have been forced to use technology to deliver their services. My physio is based down near Northampton. I’ve never met them; it’s all done online. That wouldn’t have happened for me because where I am up in the North-West, there just isn’t the services available*.” 04, stroke survivor

### Perceptions of technology

All participants were positive about the prospect of technology which could be used to support rehabilitation. Stroke survivors, particularly, saw technology as an opportunity to receive guidance when they were unable to access other services:

*“If we have the app when we leave hospital, […] he can be engaged with physio straightaway rather than being left for 2-3 years with nothing*.*”* Carer of 05

However, both stroke survivors and therapists had reservations. Therapists suggested that some survivors, particularly those with cognitive issues or perceptual problems, may not have skills to access technology-based therapy. They suggested that markerless motion capture technology could be appropriate for use with 10-20% of the stroke survivors they worked with:

*“You’re definitely not going to give it to all your patients, I think it’s going to be appropriate for maybe 20% of the caseload” A, physiotherapist*

Stroke survivors and therapists raised access to equipment as a significant barrier for uptake, often associated with cost and financial limitations:

*“We found in the pandemic that a lot of our patients can’t access technology”* A, physiotherapist

Therapists noted that users would need to be self-motivated to engage with unsupervised rehabilitation:

*“[It’s going to be] patient led. You’ve got to make sure that the patient’s on board”* B, physiotherapist

While stroke survivors were concerned that technology would not provide the same level of tailored support, or the social opportunity, of an in-person therapist.

*“But I’m not sure whether it could actually take the place of somebody being physically there*.*” 05, stroke survivor*

### Role of markerless technology for providing feedback

Physiotherapists perceived that markerless motion capture technology which provides biofeedback could provide personalised care which could respond to stroke survivors’ needs. The potential to receive user-specific guidance and feedback about quality of movement in real time was considered to be useful for stroke survivors’ development:

*“Initially looking at that, that looks amazing: the amount of data that you’re collecting, the amount of feedback you’re getting, the actual fact that the patient themselves is getting that visual feedback and commodity. There’s so many great things about it*” C, physiotherapist

Having a record of development was also suggested to be “motivational and helpful”(C) for stroke survivors and was thought likely to promote adherence.

*“The differences are so small that the patients sometimes don’t see it and then lose hope, but if they can focus on ‘right today I’ve got to achieve this’ or something like that, I think it’s a good easy goal to be recognized*.*” B, physiotherapist*

Physiotherapists mentioned specific things that needed to be considered to make any technology suitable for use with stroke survivors, particularly consideration of patient safety during use:

*“How long do we know they can tolerate it before fatigue sets in? Is that monitored via the technology[…]?” D, physiotherapist*

Physiotherapists suggested that fatigue could be monitored through movement measurement and described a need for a safety feature which stops exercises and promotes rest when user activity reaches a certain threshold.

Stroke rehab exercises are varied and affect different parts of the body, and so further concerns were raised around the range of exercises that the technology would need to include:

*“My main concern would be that they need a massive library of exercises” A, physiotherapist*

In particular, physiotherapists stressed that any technology would need to be able to be moved between positions to capture different parts of the body, and need to measure around objects used in exercises or assistive devices.

Finally, physiotherapists stressed the need for any stroke technology to be easy to use and understand, and be accessible to stroke survivor needs which can include physical difficulties, problems with communication, visual limitations, and fatigue.

*“Sometimes you might just want [guidance] to be given very very directly, using a minimal number of words*.*” C, physiotherapist*

## Discussion

As technology progresses and telerehabilitation is increasingly integrated into healthcare, it is important to consider stakeholders, their needs, and the context in which the technology will sit. This is important for supporting the development of technology for future use and ensuring that it meets the needs of the users. This study hopes to develop an intervention for stroke rehabilitation in the community, investigating how markerless motion capture technology can be adapted for community stroke rehabilitation. Participants identified positives of this technology and suggested adaptions needed for it to be useful in this setting.

The focus groups drew attention to difficulties faced by stroke survivors as they try to access sufficient rehabilitation, and some of the physical consequences of this lack of access. This is not a new finding[5] and highlights the ongoing challenges in accessing stroke rehabilitation services since the National Stroke strategy was published in 2007[27]. However, this study draws attention to the continued presence of this deficit, and its exacerbation by the Covid-19 pandemic. Insufficient access to rehabilitation can affect stroke survivors’ recovery which further negatively impacts quality of life and participation in society. Whilst technology may be used to augment existing rehabilitation services, further work is needed to ensure they are matched appropriately to the patient and that clinicians feel that this can be integrated into their practice. This is particularly important given the new guidelines which have increased the recommended dosage to 3 hours of multidisciplinary therapy a day at least 5 days a week[2]. In light of these increases, it is important to develop systems which can support this need for increased rehabilitation without significantly increasing the demand on already stretched services or further embed existing health inequalities[35]. Markerless motion capture technology could improve the quality of therapist contact time by enhancing targeted therapy and reducing contact time and treatment costs.

While the technology has not been developed for stroke survivor use at this point – participants were shown an existing system used for musculoskeletal and occupational health assessments -all participants, stroke survivors and therapists, were positive about markerless motion capture, although they were clear that, even with further development, this is not a system that will help all survivors. Participating physiotherapists suggested this kind of technology may only be useful for an estimated 10-20% of stroke survivors due to problems with cognition, perception or skills. There will also be limitations where stroke survivors or physiotherapists are not comfortable with, or cannot access, the necessary technology. However, for those able to access it, technology such as this could offer a route to improve physical rehabilitation and quality of life. Additionally, this could potentially allow for therapist resources to be reallocated or enable patients to engage with rehabilitation for longer either within or following discharge from existing services.

The Covid-19 pandemic has led to an increased use of technology and telerehabilitation within stroke rehabilitation. However, existing telerehabilitation services within the literature are focused on communication, therapist observation of movements and cognition rather than home or community-based measurement systems that provide biofeedback as a part of rehabilitation. Several other commercial based products that measure physiological parameters and provide some forms of biofeedback are available e.g. heart rate or activity monitors, however these are not integrative or specific to the range of impairments that can affect individual stroke survivors. It is important that biofeedback is context specific, considers the environment in which the action is being undertaken and delivered at a time, and in a format that is most helpful to the person undertaking rehabilitation. Research during the pandemic agreed that technology and telerehabilitation was an effective way to increase the amount of rehabilitation available to stroke survivors, providing that patients met necessary conditions to use the technology [35,36). This is further reinforced by pre-pandemic research[8,14,18,28] and highlights an area which, with further development, could increase the availability of services to stroke survivors and support wider development of resilient healthcare services. Within this study, participants were all positive about the integration of technology into community stroke rehabilitation and, providing it was developed for stroke survivors, considered it a good way to increase access to stroke rehabilitation services. Technology which uses markerless motion capture technology and accurately assesses user movement during exercises could be an easy way to facilitate telerehabilitation in the home. Whilst telerehabilitation enables increased access through removal of geographical barriers, availability of equipment, possibly driven by cost and digital provision may present and alternative barrier to both patients and services[20]. Single device markerless hardware that allows for extraction of 3D measurements is becoming more readily available in personal computing devices and mobile phones which are more readily affordable. When combined with relevant softwares, the ‘markerless systems’ could allow stroke survivors to access support without the difficulties of having to set up and manage technical equipment, whilst still collecting biomechanical data on their movements and the quality of their exercise. These systems could also be integrated into the existing care pathways in which the cost is not passed onto the user.

Participants suggested a series of necessary elements to make the presented markerless motion capture technology appropriate for stroke rehabilitation. Stroke rehabilitation is varied and so the technology needs to include a range of exercises, and must also be able to work around objects used in exercises or assistive devices. Safety should also be considered and incorporated to ensure survivors are not at risk while exercising[12]. Current rehabilitation exercises consider what the stroke survivor will be able to do alone with minimal risk of harm and this should be integrated into technology, considering risk of falls and whether the survivor is alone or accompanied when presenting exercises to the user. Physiotherapists suggested that measuring metrics like patient wellness/perception of activity and assessment of fatigue over time could form part of an exercise cut-off system to ensure patients are safe to use the technology unsupervised.

User engagement and motivation is necessary for stroke rehabilitation[37,38] and is important in situations where stroke survivors are working independently. Although engagement with rehabilitation is complex and multifaceted[38,39], incorporating elements like games, achievement, and progress tracking may help adherence. In this study, one participant mentioned how activities and monitoring progress seen in existing strength sensor-based biofeedback systems made rehabilitation fun and improved motivation to continue. Remaining in connection with a physiotherapist during exercises has also been identified as a motivator[39] and was described as a desirable feature of rehabilitation technology by stroke survivors.

Accessibility is very important. Stroke survivors have a complex range of accessibility needs which will vary between each user, and so any technology must include a range of accessibility functions. In addition to visual and audio accessibility structures already used, for example within the NHS, stroke specific needs should be considered in the construction of the technology interface and in any communication, for example the delivery of feedback and guidance to stroke survivors users [40].

Cost was regularly mentioned by all participants, including cost of the technology/app, the device to host it, and the resources and internet infrastructure to support it, similarly identified in earlier studies[8,29]. Other elements to be considered include the need for the service to last beyond the usable life of the technological device it is hosted on. Technology is developing at an intense rate, so it is important to ensure that any system built for stroke rehabilitation is integrative with other services and can remain useful, not made redundant as technology develops. A model with integrated updates for developing hardware will ensure that the technology remains relevant and useful for stroke survivors into the future. Wherever possible, effort should be made to utilise the most affordable and accessible materials possible to minimise potential users from being ostracised from healthcare services. Further considerations of aspects like integration with existing patient information systems would be useful, to ensure that records can be updated in light of training – which will improve ease of use in clinical environments.

Finally, while socialising is not the primary function of rehabilitation, the impact of loss of social links, including visits from clinicians, can impact on survivors’ wellbeing and so should continue to be considered while developing stroke services.

### Limitations

The study was impacted by Covid-19 restrictions and this significantly affected our ability to recruit; funding limitations meant that we were unable to return to this study following the pandemic. While we received some interest from stroke survivors, recruitment from therapists was limited and not very diverse. It would have been beneficial to include a wider range of therapists, particularly those from different backgrounds, to gain additional insight into their understanding of post-stroke rehabilitation. Low responses were understandable given the circumstances, but those who participated were able to give an informed response and talked openly and in detail about their experiences, discussions with the participants have offered valuable insight into areas which merit further investigation and support, particularly regarding stroke survivors’ ability to navigate the healthcare system.

While we identified personal motivation as a key theme in these focus groups, we acknowledge that there was a sampling bias in favour of stroke survivors and therapists who were already motivated enough to engage with research, their own rehabilitation, and were already comfortable using technology for video calls (and who have the finances and resources accessible for personal use). Our use of a private service mailing list for recruitment is likely have contributed towards this unbalance. However, we found that virtual recruitment and participation was beneficial for recruiting stroke survivors as it allowed us to identify and talk to individuals from across the country, some of whom said they would have been unable to participate had we conducted the focus groups in person. Virtual meetings also reduced the burden of participating for those who may have struggled to attend in person, either due to the demands of travel or the energy cost associated with participating, but we acknowledge that doing so excludes people without access to devices or the internet etc. We cannot guarantee that every stroke survivor will be as motivated to support their own rehabilitation or will have the resources to do so. Future research is needed to better understand the perceptions of people who are less willing to engage with rehabilitation or technology to explore their perceptions of markerless technology.

### Conclusion

From these focus groups, it is apparent that there is a need for support with stroke rehabilitation in the community, rehabilitation technology could provide a means to aid stroke survivors and therapists in these areas. The changes in the pandemic have increased the use of technology in healthcare. At the same time, developing technologies have improved and affordable access has increased to the public. Stroke survivors and therapists discussed problems with stroke survivors accessing services but were positive about the prospect of incorporating technology into their future, particularly where it could supplement services that were inaccessible either due to cost, availability or location. Markerless technology has the potential to be put into homes where it could provide support for stroke survivors who may be unable to access any other support and could improve the quality of therapist contact time by enhancing targeted therapy, reducing contact time and treatment costs. This could positively affect stroke rehabilitation and support stroke survivors quality of life. While care needs to be taken to ensure that the technology is accessible to all users, as the country moves into the post-pandemic era and works to further integrate technology into the healthcare pathway[24, 34], telerehabilitation which utilises markerless motion capture technology could be developed to support stroke rehabilitation services and empower stroke survivors in their rehabilitation, ensuring that they have a means to access rehabilitation continuously upon discharge from hospital. For therapists, this form of telerehabilitation could also provide an additional tool that can be used to support stroke survivors in the community.

## Data Availability

Data in the present study are not available.

## Declarations

### Ethics approval and consent to participate

This study received a favourable ethical opinion from Keele University’s Faculty of Medicine and Health Sciences Research Ethics Committee (MH-210173).

### Consent for publication

Participants consented for the use of their words in the analysis and dissemination of this research.

### Availability of data and materials

The datasets generated and/or analysed during the current study are not publicly available as participants did not give consent for their data to be shared in this way.

### Authors’ contributions

AFN was involved in the methodology, data collection, analysis, and writing of the paper. FP was involved with conceptualisation of the study, analysis and writing of the paper. EL was involved with the analysis and writing of the paper. BBH was involved with the data collection, analysis and writing of the paper. AP was a lay member of the study team and was involved with the conceptualisation of the study, data collection, analysis and writing of the paper. All authors read and approved the final manuscript.

## References

1. Lawrence, E.S., et al., Estimates of the prevalence of acute stroke impairments and disability in a multiethnic population. Stroke, 2001. 32(6): p. 1279–1284.

2. National Clinical Guideline for Stroke for the UK and Ireland. London: Intercollegiate Stroke Working Party; 2023 May 4. Available at https://www.strokeguideline.org.

3. Shumway-Cook, A. and M.H. Woollacott, Motor control: translating research into clinical practice. 2007: Lippincott Williams & Wilkins.

4. French, B., et al., Repetitive task training for improving functional ability after stroke. Cochrane database of systematic reviews, 2016(11).

5. Rozevink, S.G., et al., Effectiveness of task-specific training using assistive devices and task-specific usual care on upper limb performance after stroke: a systematic review and meta-analysis. Disability and Rehabilitation: Assistive Technology, 2021: p. 1–14.

6. Sentinel Stroke National Audit Programme (SSNAP). Clinical audit Aut-Nov 2016 (Public Report). 2016.

7. Marwaa, M.N., et al., Physiotherapists’ and occupational therapists’ perspectives on information and communication technology in stroke rehabilitation. Plos one, 2020. 15(8): p. e0236831.

8. Marwaa, M.N., C. Ytterberg, and S. Guidetti, Significant others’ perspectives on person-centred information and communication technology in stroke rehabilitation–a grounded theory study. Disability and rehabilitation, 2020. 42(15): p. 2115–2122.

9. Hayward, K.S. and S.G. Brauer, Dose of arm activity training during acute and subacute rehabilitation post stroke: a systematic review of the literature. Clinical rehabilitation, 2015. 29(12): p. 1234–1243.

10. Stockley, R., et al., Current therapy for the upper limb after stroke: a cross-sectional survey of UK therapists. BMJ open, 2019. 9(9): p. e030262.

11. Holden, M.K., T.A. Dyar, and L. Dayan-Cimadoro, Telerehabilitation using a virtual environment improves upper extremity function in patients with stroke. IEEE transactions on neural systems and rehabilitation engineering, 2007. 15(1): p. 36–42.

12. Edgar, M.C., et al., Telerehabilitation in stroke recovery: a survey on access and willingness to use low-cost consumer technologies. Telemedicine and e-Health, 2017. 23(5): p. 421–429.

13. Teasell, R., et al., Time to rethink long-term rehabilitation management of stroke patients. Topics in stroke rehabilitation, 2012. 19(6): p. 457–462.

14. Laver, K.E., et al., Telerehabilitation services for stroke. Cochrane Database of Systematic Reviews, 2020(1).

15. Allen, L., et al., Community stroke rehabilitation teams: providing home-based stroke rehabilitation in Ontario, Canada. Canadian Journal of Neurological Sciences, 2014. 41(6): p. 697–703.

16. Laver, K. and K. Osborne, Telerehabilitation in Stroke, in Telerehabilitation. 2022, Elsevier. p. 43–57.

17. Leochico, C.F.D. and N. Tyagi, Teleoccupational Therapy, in Telerehabilitation. 2022, Elsevier. p. 297–308.

18. Kerr, A., et al., Adoption of stroke rehabilitation technologies by the user community: Qualitative study. JMIR rehabilitation and assistive technologies, 2018. 5(2): p. e9219.

19. Scott B, Seyres M, Philp F, Chadwick EK, Blana D. Healthcare applications of single camera markerless motion capture: a scoping review. PeerJ. 2022 May 26;10:e13517.

20. Philp F, Faux-Nightingale A, Bateman J, Clark H, Johnson O, Klaire V, Nevill A, Parry E, Warren K, Pandyan A, Singh BM. Observational cross-sectional study of the association of poor broadband provision with demographic and health outcomes: the Wolverhampton Digital ENablement (WODEN) programme. BMJ open. 2022 Nov 1;12(11):e065709.

21. Alderwick, H. and J. Dixon, The NHS long term plan. 2019, British Medical Journal Publishing Group.

22. Burns, S.P., et al., Stroke Recovery During the COVID-19 Pandemic: A Position Paper on Recommendations for Rehabilitation. Archives of physical medicine and rehabilitation, 2022.

23. Salawu, A., et al., A proposal for multidisciplinary tele-rehabilitation in the assessment and rehabilitation of COVID-19 survivors. International journal of environmental research and public health, 2020. 17(13): p. 4890.

24. Fuentes, B., et al., Impact of the COVID-19 pandemic on the organisation of stroke care. Madrid Stroke Care Plan. Neurología (English Edition), 2020. 35(6): p. 363–371.

25. Department of Health and Social Care, U.G. A plan for digital health and social care. 2022 [cited 2022 26/08/2022].

26. Thorne, S., S.R. Kirkham, and J. MacDonald-Emes, Interpretive description: a noncategorical qualitative alternative for developing nursing knowledge. Research in nursing & health, 1997. 20(2): p. 169–177.

27. Paynter, C., et al., How people living with motor neurone disease and their carers experience healthcare decision making: a qualitative exploration. Disability and Rehabilitation, 2022. 44(13): p. 3095–3103.

28. Madsen, L.S., J. Jeppesen, and C. Handberg, “Understanding my ALS”. Experiences and reflections of persons with amyotrophic lateral sclerosis and relatives on participation in peer group rehabilitation. Disability and Rehabilitation, 2019. 41(12): p. 1410–1418.

29. Bright, F.A., C.M. McCann, and N.M. Kayes, Recalibrating hope: A longitudinal study of the experiences of people with aphasia after stroke. Scandinavian Journal of Caring Sciences, 2020. 34(2): p. 428–435.

30. Im, J., et al., “Whatever happens, happens” challenges of end-of-life communication from the perspective of older adults and family caregivers: a Qualitative study. BMC palliative care, 2019. 18(1): p. 1–9.

31. Boland, L., D.I. McIsaac, and M.L. Lawson, Barriers to and facilitators of implementing shared decision making and decision support in a paediatric hospital: a descriptive study. Paediatrics & child health, 2016. 21(3): p. e17–e21.

32. Tong, A., P. Sainsbury, and J. Craig, Consolidated criteria for reporting qualitative research (COREQ): a 32-item checklist for interviews and focus groups. International journal for quality in health care, 2007. 19(6): p. 349–357.

33. Malterud, K., Siersma, V. D., & Guassora, A. D. (2016). Sample size in qualitative interview studies: guided by information power. Qualitative health research, 26(13), 1753–1760.

34. Clarke, V. and V. Braun, Thematic analysis: a practical guide. Thematic Analysis, 2021: p. 1–100.

35. Ostrowska PM, Śliwiński M, Studnicki R, Hansdorfer-Korzon R. Telerehabilitation of post-stroke patients as a therapeutic solution in the era of the COVID-19 pandemic. InHealthcare 2021 May 31 (Vol. 9, No. 6, p. 654). MDPI.

36. Silakarma D, Widnyana M. Telerehabilitation as a physical therapy solution for the post-stroke patient in COVID-19 pandemic situations: A review I Made Yoga Prabawa. Intisari Sains Medis. Intisari Sains Medis. 2021;12:1–5.

37. Lequerica AH, Kortte K. Therapeutic engagement: a proposed model of engagement in medical rehabilitation. American journal of physical medicine & rehabilitation. 2010 May 1;89(5):415–22.

38. MacDonald GA, Kayes NM, Bright F. Barriers and facilitators to engagement in rehabilitation for people with stroke: a review of the literature. New Zealand Journal of Physiotherapy. 2013 Nov 1;41(3).

39. Forgea MC, Lyons AG, Lorenz RA. Barriers and facilitators to engagement in rehabilitation among stroke survivors: an integrative review. Rehabilitation Nursing Journal. 2021 Nov 1;46(6):340–7.

40. Stroke Foundation. Communication after Stroke. 2024.https://strokefoundation.org.au/what-we-do/for-survivors-and-carers/after-stroke-factsheets/communication-after-stroke-fact-sheet. Last accessed on 29th January 2024.

