## Appendix 1 for "Exploring stroke survivors’ and physiotherapists’ perspectives of the potential for markerless motion capture technology in community rehabilitation"

**Appendix 1 - Focus Group Schedule**

**Stroke survivor focus group**

- We’d like to know about the transition from hospital-based to community-based rehab.
  - E.g. Were you given any advice when you were discharged? Did you have a good understanding of what that meant for you?
  - Do you access any services for your rehabilitation?
  - What are you currently doing at home (has rehabilitation changed due to the pandemic?)
- What motivates you to engage or not engage with your exercises?
- What are your thoughts about stroke technology, particularly markerless motion capture technology (explanation)? Do you think that this sounds like something that you might find useful?
- What factors would influence your use of technology?
- Are there any elements that you can think of that would be important to consider when developing stroke technology?
  - E.g. What support would you like technology to provide?
  - Are there any requirements that you have? E.g. ease of use – single button/easy to use interface, large print etc.

**Therapist focus group**

- How do you find that stroke survivors respond to rehabilitation?
  - E.g. What are your experiences of stroke rehabilitation services?
  - How are services currently running?
- What are your thoughts about stroke technology, particularly markerless motion capture technology (explanation)? Could technology support the treatment you currently offer to stroke patients?
  - Seed questions: Could you use technology in the hospital as part of your current regime and then encourage patients to continue use with it at home?
  - Could it bridge the gap between these locations?
- If it were available, would you use technology like markerless motion capture technology in your stroke rehabilitation service? Why?
- What would technology need to have to be useful to you and the patients you work with?
  - Seed questions: would you want measurements/tests across intervals as a measure of participation and marker of improvement?
- Are there any additional requirements/ideas you have that you would be interested in?
- What do you think a patient can do within constraints of home environment
- How do you customise the exercises for them? E.g use a can of beans? Got to make sure it doesn’t affect measuring the movements
